# The maternal profile associated with the underlying root cause of neonatal mortality using machine learning and administrative health data

**DOI:** 10.1101/2022.08.15.22278783

**Authors:** Carlos Eduardo Beluzo, Tiago Carvalho, Luciana Correia Alves

## Abstract

Infant mortality is a reflection of an analysis of biological, socioeconomic, and assistance factors. An analytical analysis of this problem implies the processing of large sets of data from different areas. Data Science approaches have become increasingly widespread to deal with problems that require large datasets to perform deep analysis. Machine learning methods have become popular due to their efficiency and efficacy in discovering knowledge by identifying patterns in feature interactions of large datasets. This work proposes the use of a machine learning approach to evaluate the association between sociodemographic factors and preventable root causes of neonatal mortality. For this, demographic and epidemiological data from Brazilian public health birth and mortality (SINASC and SIM, respectively) information systems were used. Using an unsupervised approach, for instance, the *K-Modes* clustering algorithm, clusters were created, so we are able to evaluate the socio-demographic profile of each one of the clusters. In this way, it is possible to evaluate the differences between the profiles of each cluster. The profile consists of features such as maternal age, maternal years of schooling, race, number of consultations, type of delivery, public or private hospital, and date of first prenatal consultation. The analysis was performed using data from the period between 2012 and 2018, for the city of São Paulo, one of the richest regions of the country. The data quality for this region is considered to be very high, so there is no need to apply data correction methods. Besides that, the method adapted does not require data assumptions, and it’s suitable for categorical data, which is our case. Considering that this is a data-driven approach, preliminary results indicate that only a few assumptions can be made on profile using these features, although some associations between demographic variables and neonatal mortality by preventable root causes can be identified. We hope to encourage reflection on the newborn in the socioeconomic environment and contribute to public health policies.

## Introduction

According to WHO definitions, **child mortality** is a survival measure that refers to children dying before reaching five years of age and is also referred to as **under-5 mortality. Infant Mortality (IM)** represents an essential component of under-5 mortality and is a measure that relates to children dying before reaching the first year of life. These deaths reflect the social, economic, and environmental conditions in which children (and others in society) live, including their health care, so they are good measures to identify vulnerable populations. The infant mortality rate is a Millennium Development Goals (MDG) indicator [1].

IM can also be disaggregated into Neonatal Mortality (NM) and Postneonatal mortality, and the first one has been an essential object of study in the last decades, especially because it concentrates on the higher share of deaths of IM. Formally, the IM rate is a probability of death derived from a life table and expressed as a rate per 1,000 live births, and the same is valid for the Neonatal Mortality rate (NMR).

In the last 65 years, the global mortality rate has declined 5 times, however, disparities between developed and developing countries demonstrate that there is work to do. The fact that mortality rates can be 10 times higher in developing countries indicates that a high percentage of these deaths can be avoidable [2].

Evaluating determinants of infant mortality can favor the better targeting of public policy funding, which is increasingly possible through online platforms that make data available, such as the Department of Brazilian Health Ministry (DATASUS) responsible for collecting, processing, and disseminating public health data [2].

In Brazil, the IMR has undergone a continual decrease in recent decades, mainly due to improvements in sanitary conditions and the reduction in Postneonatal which has declined from 51% in 1990 to 38% in 2015 [3]. Nowadays the NM which is the main component of the IMR has also been following a declining trend, mainly due to favorable changes in factors related to pregnancy and childbirth, despite the increase in its shared proportion [2].

Neonatal mortality involves various biological, socioeconomic, and health-care factors and has become the biggest challenge in fighting infant mortality. The increasing importance of neonatal deaths on infant mortality aside from the greater availability and quality of health data in Brazil has enabled more precise analyses and led to a significant number of studies covering different factors, regions, and methods regarding this issue [2].

In this paper, it is proposed an evaluation of the NM in the city of São Paulo - Brazil, in the period between 2012 and 2018, considering maternal and socio-demographic information available from public health records, and preventable underlying causes of death. According to data from the Brazilian Mortality Information System (SIM), there were 13,309 deaths of under-5 children in this period in the municipality of São Paulo. **Figure 1** depicts these numbers desegregated by stage of the child’s life, from every 8,180 deaths refers to neonatal deaths.

**Figure 1.**
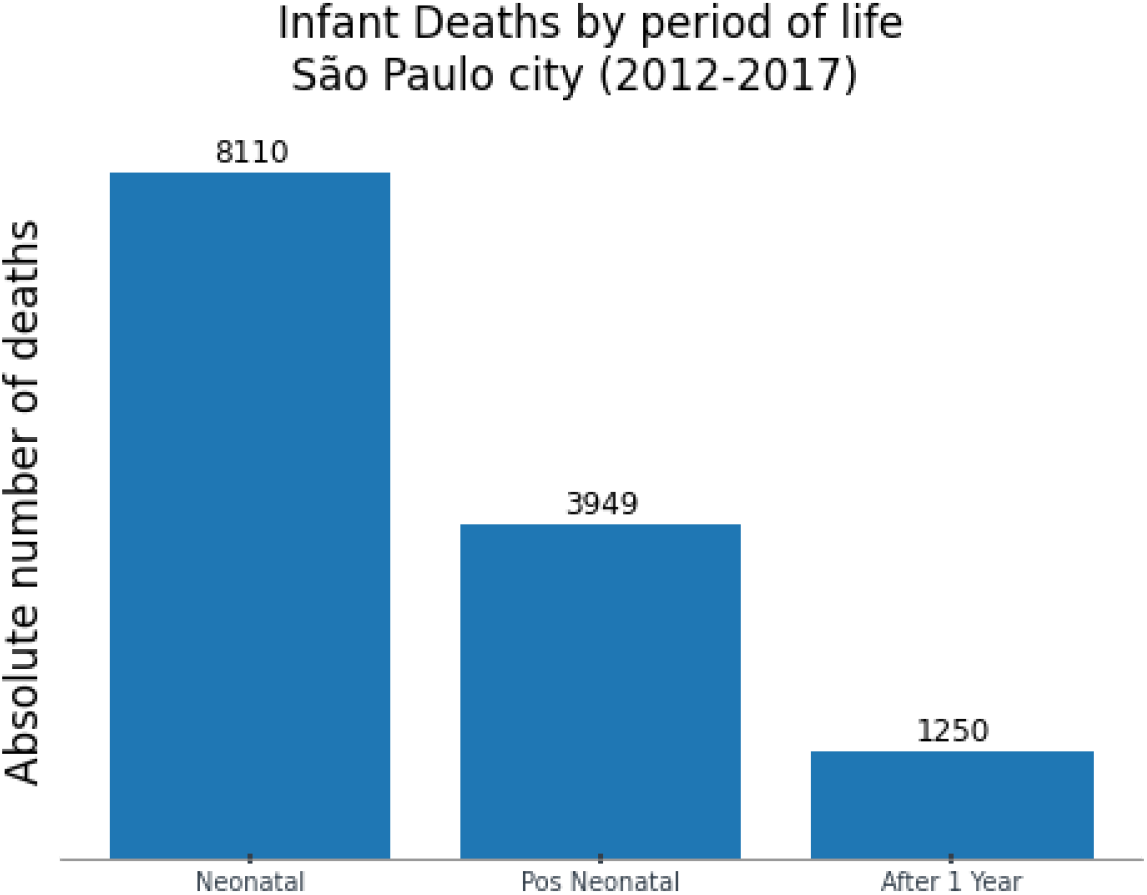
*Infant deaths distributed by mortality components in the city of São Paulo between 2012 and 2018.* (Source: SIM-DATASUS)

## Dataset

The data came from Mortality Information System (SIM - Sistema de Informação de Mortalidade) and the National Information System on Live Births (SINASC – Sistema de Informação de Nascidos Vivos), both from DATASUS (Health Informatics Department of the Brazilian Ministry of Health). From these, it was extracted the dataset used in this paper, which has been built by selecting only records from São Paulo city for the period of 2012 to 2018. Data from São Paulo city has high-level quality. Although São Paulo has one of the lowest levels of neonatal mortality rates in Brazil, these events occurred in heterogeneous ways with smaller or even lower reductions in the most vulnerable populations, as the reflection of unfavorable life conditions of the population, healthcare and socioeconomic inequalities [5].

SINASC is fed using the Live Birth Statement (DNV - Declaração de Nascido Vivo). It comprises information about demographic and epidemiological data for the infant, mother, prenatal care, and childbirth. On the other hand, SIM has the main goal of supporting the collection, storage, and management process of death records in Brazil, and was used to label records where the death happened until 28 days of life on SINASC, using DNV, which is a common field in both systems, as an association key [6]. Methodology used to create this dataset was the same used to build the **SPNeodeath** dataset [7]. **Table 1** depicts the selected features for the current experiment.

**Table 1.**
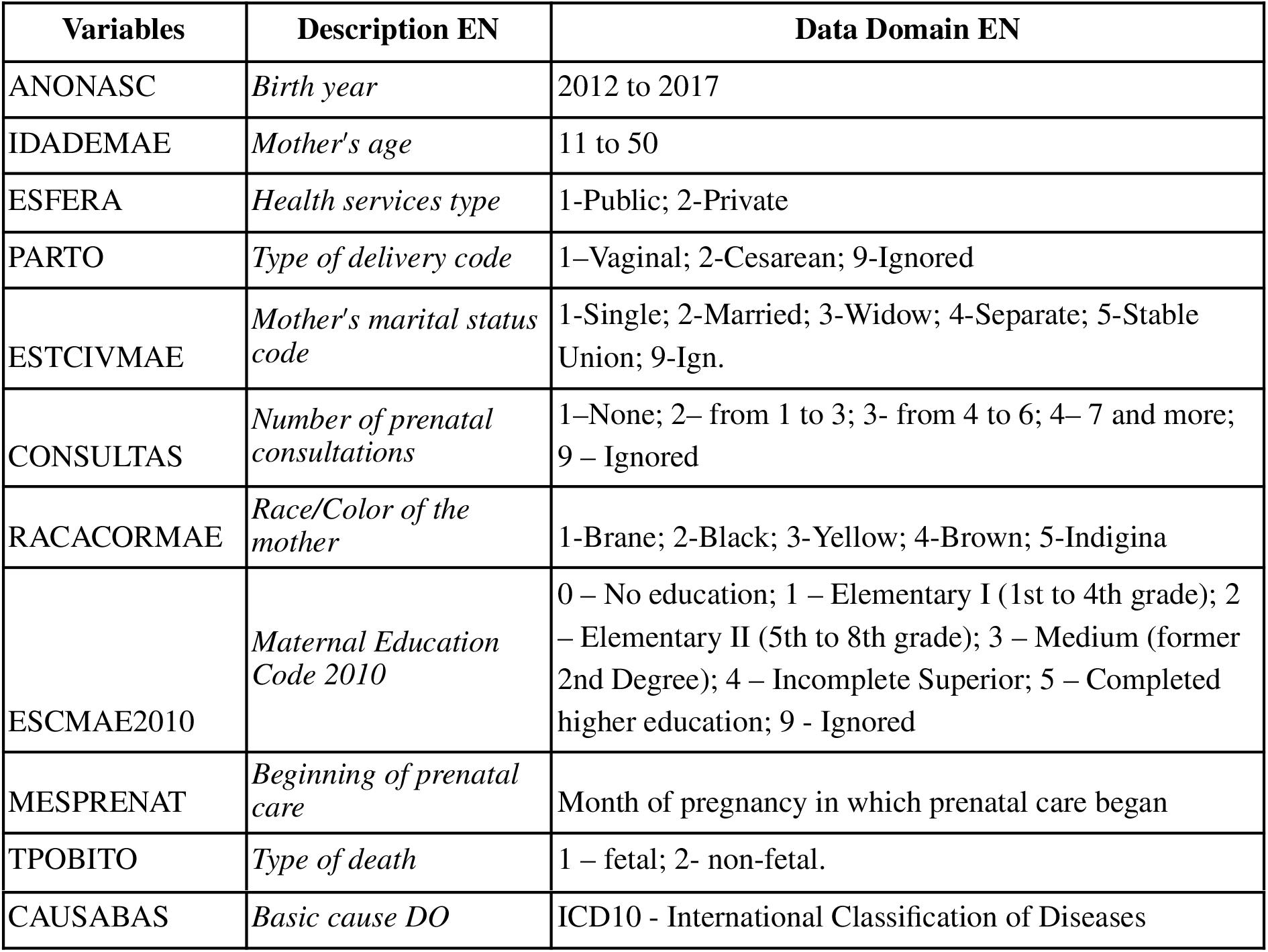
Data dictionary of the SIM and SINASC features selected to be used in this paper.

Currently, the underlying causes of death were classified into three groups *[4]*:

a. *avoidable causes*;
b. *Ill-defined causes of death, and*;
c. *Other causes of death (not clearly preventable)*.

One of the common approaches in mortality studies is to evaluate the root cause of death, and it will be part of the object of study of this paper. Among the various root causes, this paper will focus on the <u>avoidable causes</u>. The causes of death of this group are in whole or in part preventable by effective actions of health services that are accessible at a given place and time. These causes must be reviewed in light of the evolution of knowledge and technologies for health care practices. Within this group there are also the following categories *[4]*:

i. r*educible by immunoprevention actions*;
ii. *reduced by adequate care for women during pregnancy, childbirth, fetus, and newborn*;
iii. *reducible by adequate diagnostic and treatment actions, and*;
iv. *reducible by adequate health promotion actions, linked to adequate health promotion actions health care*.

### Proposed method

The main objective is to perform a clustering of neonatal deaths to compare differences in sociodemographic and maternal profiles, which could be associated with preventable underlying causes of death. When choosing the method, it was established that no data assumption will be taken regarding the data, except that all selected features are categorical. These requirements lead to the algorithm being adopted as a feasible option. So, in this paper, an unsupervised Machine Learning approach is being used to build a maternal and socio-demographic profile through a clustering approach.

Specifically, the *K-mode Clustering* algorithm was adopted, once it is an algorithm that represents a natural formulation of clustering for categorical data, in a general way, it extends the *k-means* paradigm to a categorical domain [8]. It defines clusters based on the number of matching categories between data points, in contrast to the more well-known *k-means* algorithm, which clusters numerical data based on Euclidean distance [9]. This paper uses the Python library implementation *kmodes* version 0.12.

As the main objective is to associate maternal and socioeconomic profiles with underlying causes of death, when executing the algorithm, this feature has been removed, thus avoiding bias. After the clusters have been created, each sample is categorized in one of them, and finally, the root cause feature is reinserted. Thus, at the end of the process, we have a dataset where it is possible to perform an analysis of association.

### Preliminary Results

The most frequent underlying causes of neonatal mortality in São Paulo between 2012 and 2018 are depicted in **Figure 2**, from which 90% are associated with 33 underlying causes (including root causes from all groups) and the top six are:

1. **P36-9**: *Bacterial sepsis of newborn, unspecified*;
2. **P22-0**: *Respiratory distress syndrome of the newborn*;
3. **P00-0**: *Fetus and newborn affected by maternal hypertensive disorders*;
4. **P01-1**: *Fetus and newborn affected by premature rupture of membranes*;
5. **P96-9**: *Condition originating in the perinatal period, unspecified, and;*
6. **P24-9**: *Neonatal aspiration syndrome, unspecified*.

**Figure 2.**
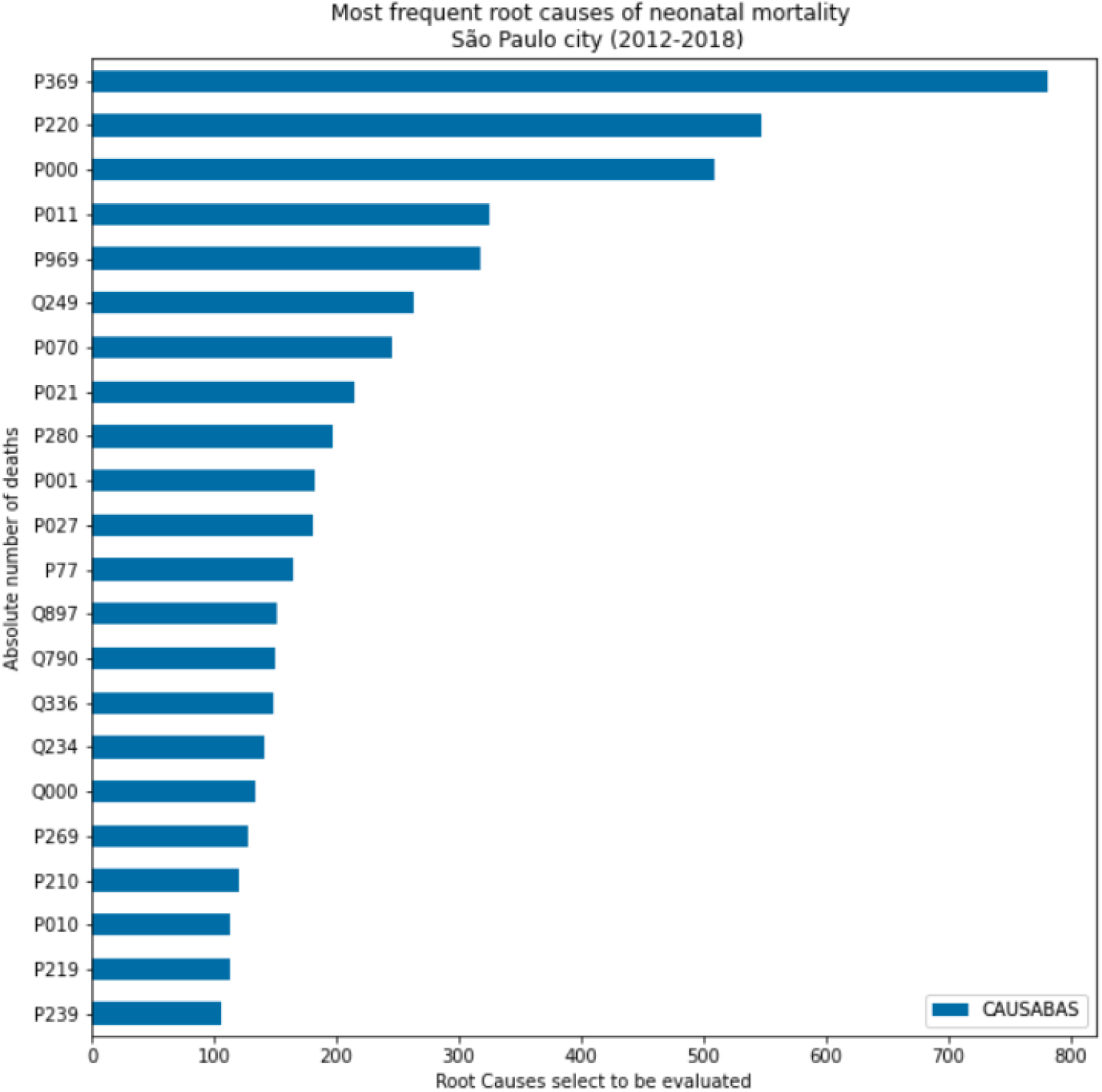
*Most frequent underlying root causes of neonatal mortality in São Paulo city between 2012 and 2018.* (Source: SIM-DATASUS)

From this result, only the root cause **P96-9** is not classified in the category (**a-ii)** (*reduced by adequate care for women during pregnancy, childbirth, fetus, and newborn*), but, as it is classified as **(b)** (*Ill-defined causes of death*), which may also be including avoidable causes.

Initially, a few rounds of experiments were carried out, varying the number of clusters using the entire dataset, but the results were inconclusive or just a random classification. So, as the first approach to improving the model, the experiment was carried out using 90% of the dataset, filtering by the most frequent root causes, in which the possible causes of death were reduced from 378 to 24. In a very optimistic approach, the algorithm was configured to create 24 clusters, and as expected, after performing a projection of each of the profiles on a scatter plot, in visual analysis, a completely random distribution is also observed.

In a slightly more coherent way, the number of clusters was reduced to 6, considering that 50% of the dataset is distributed in 6 root causes, however, by a simple visual analysis of the scatter plots in **Figure 3**, we cannot conclude that there is any association with the profiles and root causes. Each of the six scatter plots in the figure represents one of the clusters created by the algorithm, and the events that were associated with it are projected on the graphic. The colors represent the root causes; the *y-axis* represents the event itself, and the *x-axis* represents each of the possible profiles. The profiles were created using a simple combinatorial mathematical process, in order to favor the visualization in two dimensions. Therefore, each point represents an individual, the position in “x” its profile, and its color, the underlying cause of death.

**Figure 3.**
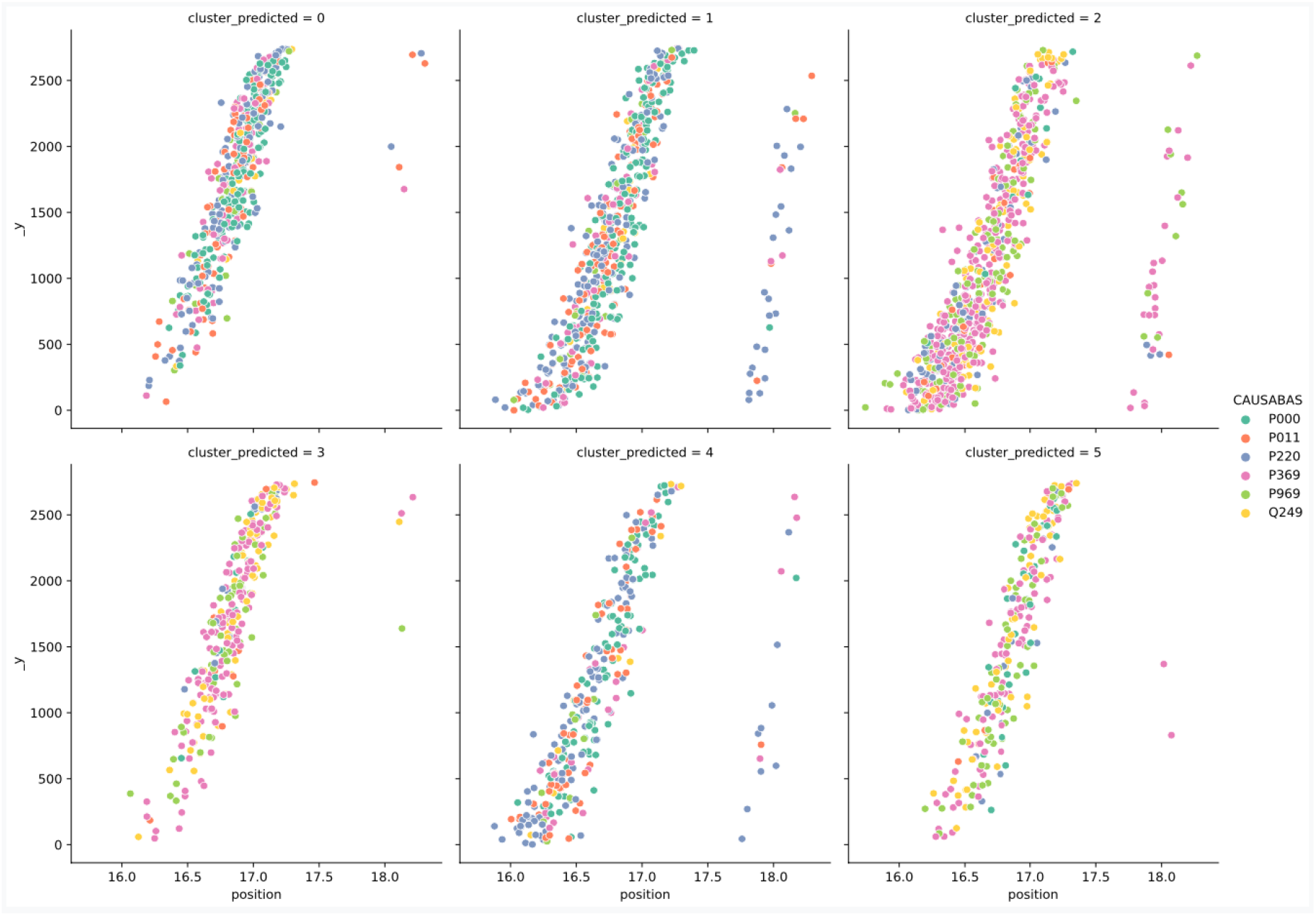
Clusters created by K-modes algorithm grouping neonatal deaths by sociodemographic profiles and associated with the underlying cause of death.

By visual analysis, it is possible to infer that the algorithm was able to group in clusters 2, 3, and 5 a higher incidence of individuals classified with the root causes **Q24-9** and **P36-9**, however, by visual inspection of the graphic nothing can be stated about the profiles since the clusters present similar random distributions among them. The experiment was also carried out by setting the classification task for only two clusters, presenting also inconclusive results regarding the profiles, as depicted in **Figure 4**.

**Figure 4.**
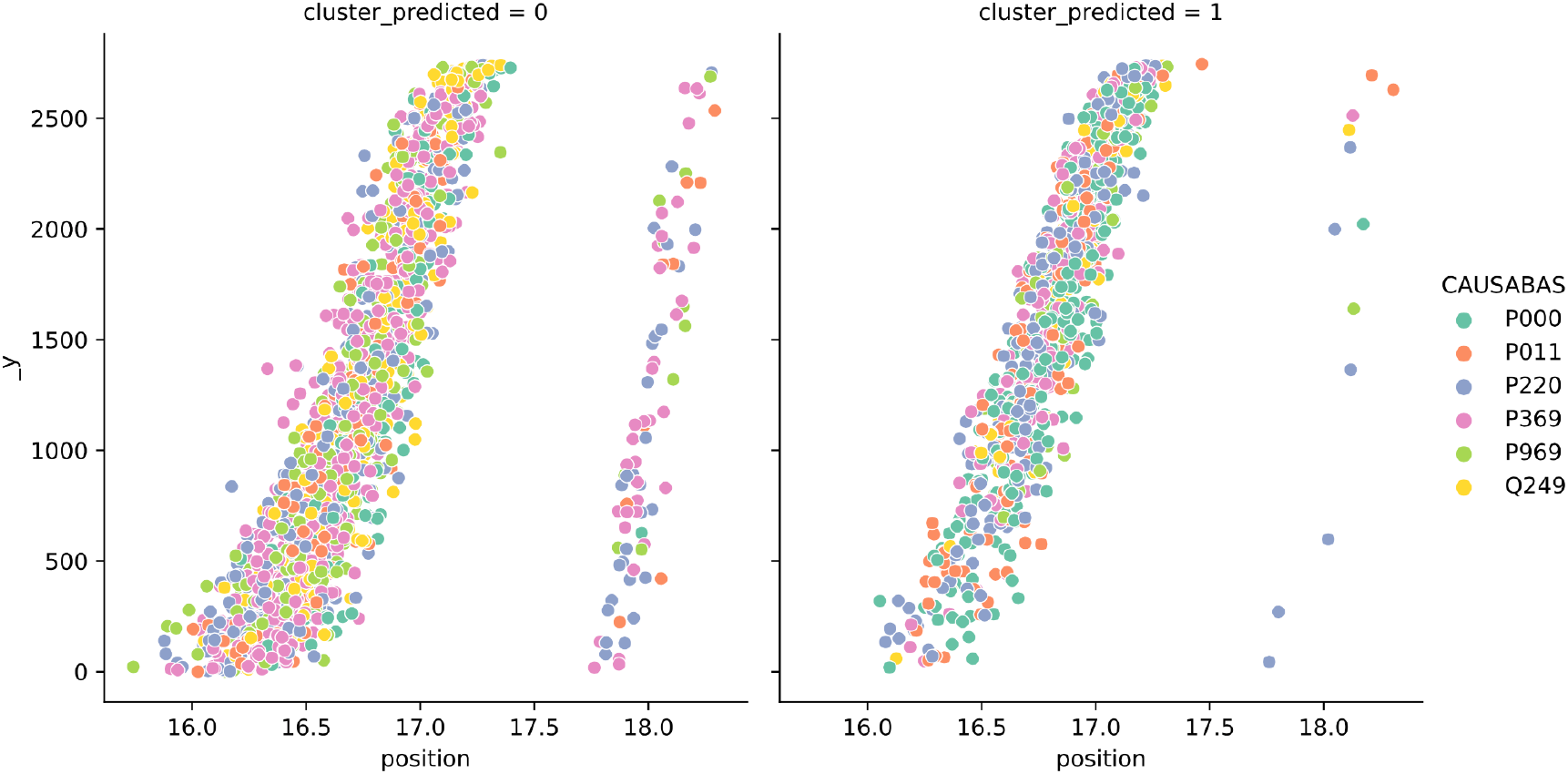
Clusters created by K-modes algorithm group neonatal deaths by sociodemographic profiles and associated with root cause death.

Thus, to date, no association can be established considering the root causes of death individually. Therefore, the second round of experiments was started, which are still under development. In this new approach, each of the events is categorized by considering whether its root cause of death corresponds to a preventable underlying cause or not. Thus, the final experiment will seek to build two clusters and assess the existence of an association between the maternal socioeconomic profile and each of the clusters, as well as the association of profiles with preventable causes.

## Data Availability

All data produced in the present study are available upon reasonable request to the authors.

https://doi.org/10.1016/j.dib.2020.106093

https://opendatasus.saude.gov.br/dataset/sim-2020-2021

https://opendatasus.saude.gov.br/dataset/sistema-de-informacao-sobre-nascidos-vivos-sinasc-1996-a-2020

